# Genetics of Smoking and Risk of Atherosclerotic Cardiovascular Diseases: A Mendelian Randomization Study

**DOI:** 10.1101/2020.04.07.20053447

**Authors:** Michael G. Levin, Derek Klarin, Themistocles L. Assimes, Matthew S. Freiberg, Erik Ingelsson, Julie Lynch, Pradeep Natarajan, Christopher O’Donnell, Daniel J. Rader, Philip S. Tsao, Kyong-Mi Chang, Benjamin F. Voight, Scott M. Damrauer, on behalf of the VA Million Veteran Program

**Affiliations:** Division of Cardiovascular Medicine, University of Pennsylvania Perelman School of Medicine, Philadelphia, PA; Department of Medicine, University of Pennsylvania Perelman School of Medicine, Philadelphia, PA; Corporal Michael J. Crescenz VA Medical Center, Philadelphia, PA; Malcolm Randall VA Medical Center, Gainesville, FL; Department of Surgery, University of Florida, Gainesville, FL; Palo Alto VA Healthcare System, Palo Alto, CA; Department of Medicine, Division of Cardiovascular Medicine, Stanford University School of Medicine, Stanford, CA; Stanford Cardiovascular Institute, Stanford University, Stanford, CA; Division of Cardiovascular Medicine, Vanderbilt University Medical Center, Nashville, Tennessee; Geriatric Research Education and Clinical Centers (GRECC), Veterans Affairs Tennessee Valley Healthcare System, Nashville, Tennessee; Department of Medicine, Vanderbilt University Medical Center, Nashville, Tennessee; Stanford Diabetes Research Center, Stanford University, Stanford, CA; Edith Nourse VA Medical Center, Bedford, MA; VA Informatics and Computing Infrastructure, Salt Lake City, UT; Cardiovascular Research Center, Massachusetts General Hospital, Boston, MA; Broad Institute of Harvard and MIT, Cambridge, MA; Department of Medicine, Harvard Medical School, Boston, MA; VA Boston Healthcare System, Boston, MA; Department of Genetics, University of Pennsylvania Perelman School of Medicine, Philadelphia, PA; Institute for Translational Medicine and Therapeutics, University of Pennsylvania Perelman School of Medicine, Philadelphia, PA; Department of Medicine, Division of Cardiovascular Medicine, and Stanford Cardiovascular Institute, Stanford University, Palo Alto, CA; Department of Systems Pharmacology and Translational Therapeutics, University of Pennsylvania Perelman School of Medicine, Philadelphia, PA; Department of Surgery, University of Pennsylvania Perelman School of Medicine, Philadelphia, PA

**Keywords:** smoking, genetics, atherosclerosis, Mendelian Randomization, cardiovascular disease, coronary artery disease, peripheral artery disease, stroke

## Abstract

**Importance:** Smoking is associated with atherosclerotic cardiovascular disease, but the relative contribution to each subtype (coronary artery disease [CAD], peripheral artery disease [PAD], and large-artery stroke) remains less well understood.

**Objective:** To determine the effect of smoking on risk of coronary artery disease, peripheral artery disease, and large-artery stroke.

**Design:** Mendelian randomization study using summary statistics from genome-wide associations of smoking (up to 462,690 individuals), coronary artery disease (up to 60,801 cases, 123,504 controls), peripheral artery disease (up to 24,009 cases, 150,983 controls), and large-artery stroke (up to 4,373 cases, 406,111 controls)

**Setting:** Population-based study of primarily European-ancestry individuals

**Participants:** Participants in genome-wide association studies of smoking, coronary artery disease, peripheral artery disease, and stroke.

**Exposures:** Genetic liability to smoking defined by lifetime smoking index: an integrated measure of smoking status, age at initiation, age at cessation, number of cigarettes smoked per day, and declining effect of smoking on health outcomes).

**Main Outcome Measure:** Risk of coronary artery disease, peripheral artery disease, and large-artery stroke.

**Results:** Genetic liability to smoking was associated with increased risk of PAD (OR 2.13; 95% CI 1.78-2.56; P = 3.6 × 10^−16^), CAD (OR 1.48; 95% CI 1.25-1.75; P = 4.4 × 10^−6^), and stroke (OR 1.4; 95% CI 1.02-1.92; P = 0.036). Risk of PAD in smokers was greater than risk of large-artery stroke (p_difference_ = 0.025) or CAD (p_difference_ = 0.0041). The effect of smoking on ASCVD remained independent from the effects of smoking on traditional cardiovascular risk factors.

**Conclusions and Relevance:** Genetic liability to smoking is a strong, causal risk factor for CAD, PAD, and stroke, although the effect of smoking is strongest for PAD. The effect of smoking is independent of traditional cardiovascular risk factors.

## INTRODUCTION

Atherosclerotic cardiovascular disease (ASCVD) can affect multiple vascular beds throughout the body, with clinical manifestations including coronary artery disease (CAD), stroke, and peripheral artery disease (PAD). Smoking tobacco is consistently among the leading risk factors for ASCVD; however, the relative contribution of smoking to the individual ASCVD outcomes remains less well studied. Observational studies have examined these ASCVD outcomes together, with a recent study by Ding et. al. finding the strongest effect of smoking on incident PAD compared to CAD or stroke.^1–4^ Observational study designs may be limited, however, by modest overall sample size, measurement error, and risk of residual confounding.^5^

A number of studies over the past several decades have identified detrimental effects of smoking on traditional cardiovascular risk factors including blood pressure, lipids, and diabetes.^6,7^ Smoking also has additional independent effects on inflammation, endothelial function, and platelet aggregation.^6^ Despite the clear observational links between smoking and atherosclerosis, whether the effect of smoking on ASCVD is primarily mediated through correlated alterations of traditional cardiovascular risk factors, or operates via independent mechanisms is less clear. As the detrimental effects of smoking may persist for decades,^4^ clarifying the basis of the smoking-atherosclerosis relationship could enable more targeted risk-reduction strategies among both current and former smokers, and identify novel treatment strategies for those at highest risk of ASCVD.

Recently, large genome-wide association studies of smoking, coronary artery disease, stroke, and peripheral artery disease have identified genetic loci associated with each of these conditions.^8–11^ The Mendelian randomization (MR) framework leverages the natural randomization of genetic variation at conception (under certain assumptions) to mitigate the risk of confounding when estimating relationships between traits. Therefore, this approach may allow for more precise quantification of potential differences between exposure-outcome pairs.^12^ The method has further been extended to consider exposures jointly, a form of mediation analysis that enables the estimation of the direct effect of each exposure on an outcome of interest.^13^

Here, leveraging population-scale human genetics data from genome-wide association studies, we utilized the MR framework with genetic variants as instrumental variables to: (i) estimate the total causal effect of smoking on coronary artery disease, peripheral artery disease, and large-artery stroke, the primary manifestations of ASCVD; (ii) quantify differences in the estimates of effects of smoking between ASCVD outcomes; (iii) validate the effect of smoking on traditional cardiovascular and inflammatory risk factors for ASCVD; and (iv) assess the extent to which traditional cardiovascular and inflammatory risk factors mediate the relationship between smoking and ASCVD outcomes.

## METHODS

### Genetic Instrument Selection

The lifetime smoking index is a continuous measure that accounts for self-reported smoking status, age at initiation, age at cessation, number of cigarettes smoked per day, and a simulated half-life constant that captures the declining effect of smoking on health outcomes following a given exposure.^11^ A genetic instrument for lifetime smoking index was constructed from summary statistics of a genome-wide association study of UK Biobank participants (UK Biobank; N = 462,690) using independent (r^2^ = 0.001, distance = 10,000 kb, European-ancestry participants of the 1000 genomes project) genome-wide significant (p < 5 × 10^−8^) variants as the exposure, as previously described.^11^ Each standard-deviation increase in the lifetime smoking index instrument corresponds to an individual smoking 20 cigarettes daily for 15 years and stopping 17 years ago, or smoking 60 cigarettes daily for 13 years and stopping 22 years ago.

The corresponding SNP effects for CAD^8^ (CARDIoGRAMplusC4D 1000 Genomes; N = 60,801 cases, 123,504 controls), PAD^9^ (Million Veterans Program; N = 24,009 cases, 150,983 controls), and large-artery stroke^10^ (MEGASTROKE; N = 4,373 cases, 406,111 controls) were used as the outcomes, without the use of proxy SNPs. The smoking index instrument was previously validated using an independent set of participants and with positive-control outcomes of coronary artery disease and lung cancer. To further validated the instrument, two-sample MR was performed using lifetime smoking index as the exposure, and smoking initiation, age of smoking initiation, smoking cessation, and cigarettes per day,^14^ as reported by GSCAN, as outcomes. To evaluate the strength of the smoking index instrument, the F-statistic was calculated.^15^

### Mendelian Randomization

In the primary analysis, the total effect of lifetime smoking index on ASCVD outcomes (CAD, PAD, and stroke) was estimated using random-effects inverse-variance weighted MR within the *TwoSampleMR* package in R.^165^ In sensitivity analyses, fixed-effects inverse-variance weighted, maximum likelihood, weighted-median, penalized weighted-median, and MR-PRESSO methods were performed to account for potential violations of the instrumental variable assumptions, heterogeneity, and error in the instrument-exposure associations.^5,17,18^ Diagnostic leave-one-out, single-SNP, funnel-plot analyses were performed. The Egger bias intercept test was used to detect evidence of horizontal pleiotropy. In sensitivity analysis, a genetic instrument for smoking initiation was used as the exposure.^14^ Because two-sample Mendelian randomization of binary exposures provides effect estimates per 1-unit change in the exposure, results of the effect of the smoking initiation exposure on ASCVD outcomes are expressed as odds of the outcome per 2.72-fold (1 log-odds unit) increase in the odds of ever smoking.^19^ For the primary analysis, power calculations were performed to estimate the minimum effect for which the study had an 80% power to detect a difference at a two-tailed significance level of p < 0.05. Differences in the effect of smoking on ASCVD outcomes in each vascular bed was estimated using a test of interaction.^20^

The effect of lifetime smoking on cardiometabolic risk factors was estimated using random-effects inverse-variance weighted two-sample MR. Outcomes were obtained from publicly available summary statistics from genome-wide association studies of continuous traits (total cholesterol, LDL-cholesterol, HDL-cholesterol, triglycerides, BMI, waist-to-hip ratio, fasting glucose, fasting insulin, systolic blood pressure, estimated glomerular filtration rate, circulating C-reactive protein levels, circulating IL1B, circulating IL6, and circulating IL6R levels), and binary traits (type II diabetes mellitus, hypertension, hyperlipidemia, chronic kidney disease, overweight) identified using the MR-Base platform **(Supplemental Table 1)**.^21–28^ Because the lifetime smoking index exposure instrument was derived from UK Biobank participants, and MR estimates derived from studies with a high proportion of overlapping samples may be biased, studies of cardiometabolic outcomes that included non-UK Biobank participants were preferred when available.^29^ The smoking initiation exposure, which included both UK Biobank participants and participants from several other studies, was also used in sensitivity analysis to further minimize bias from participant overlap. The MR Steiger test of directionality was performed to validate direction of association between smoking (as the exposure) and cardiometabolic risk factors (as the outcomes).^30^

To determine whether any effect of smoking on ASCVD may be mediated through traditional cardiovascular risk factors, multivariable MR was performed.^31,32^ Independent (r^2^ = 0.001, distance = 10,000 kb) genome-wide significant (p < 5 × 10^−8^) variants associated with traditional cardiovascular and inflammatory risk factors (LDL-cholesterol, HDL-cholesterol, triglycerides, BMI, type II diabetes mellitus, systolic blood pressure, and circulating IL6 levels) were identified using the MR-Base platform. The direct effect of lifetime smoking index was then estimated in models accounting for each traditional risk factor alone, and in a model considering all risk factors jointly.

### Statistical Analysis

For the primary analysis of smoking on ASCVD outcomes, the statistical significance threshold was set at a two-sided p < 0.05. For the secondary analysis of smoking on cardiometabolic risk factors, correction for multiple comparisons was made using the Bonferroni method (p < 0.05/21 = 0.0024). Cardiometabolic traits with nominal associations (0.0024 ≤ p < 0.05) were considered suggestive. In the multivariable analysis considering joint effects of smoking and risk factors on ASCVD outcomes, the statistical significance threshold was set at a two-sided p < 0.05. All analyses were performed using R version 3.6.2 (R Foundation for Statistical Computing).

## RESULTS

### Selection of Genetic Variants as Proxies for Smoking

Of the 126 independent SNPs associated with lifetime smoking index, 116, 107, and 105 were available in the stroke, CAD, and PAD summary statistics **(Supplemental Table 2)**. For the lifetime smoking index genetic instrument, the F-statistic was greater than 10 (range 25-163, mean 41), suggesting low risk of weak-instrument bias. To validate the genetic instrument for lifetime smoking index, MR was performed using smoking traits from the GSCAN consortium as outcomes. Lifetime smoking index significantly increased risk of smoking initiation and cessation, increased amount of smoking (cigarettes per day), and decreased age of smoking initiation **(Supplemental Figure 1)**.

**Figure 1:**
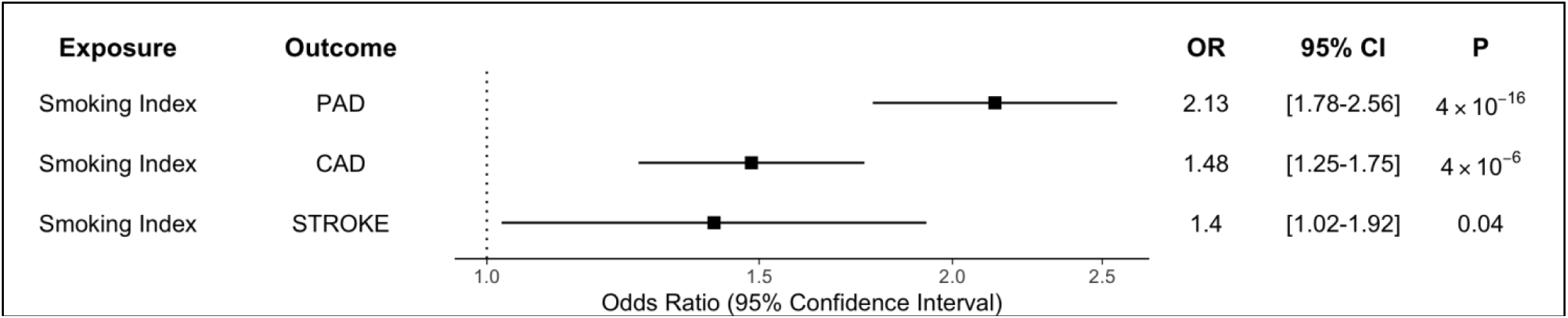
Total effect of smoking on risk of peripheral artery disease (PAD), coronary artery disease (CAD), and stroke. In inverse-variance weighted MR, each one standard deviation increase in genetic liability to smoking was associated with significantly increased risk of PAD, CAD, and large-artery stroke. Smoking most strongly increased risk of PAD compared to large-artery stroke (p_difference_ = 0.025) and CAD (p_difference_ = 0.0041). Odds ratios are expressed per one standard deviation increase in lifetime smoking index. OR = Odds Ratio, CI = Confidence Interval.

### Effect of Smoking on Atherosclerotic Cardiovascular Disease Outcomes

For the primary analysis, power calculations indicated the lifetime smoking index instrument provided >80% power to detect ORs >1.15 for CAD, >1.22 for PAD, and >1.52 for large-artery stroke. In inverse-variance weighted Mendelian randomization analysis, each 1 standard deviation increase in genetic liability to lifetime smoking index increased risk of PAD (OR 2.13; 95% CI 1.78-2.56; P = 3.6 × 10^−16^), CAD (OR 1.48; 95% CI 1.25-1.75; P = 4.4 × 10^−6^), and stroke (OR 1.4; 95% CI 1.02-1.92; P = 0.036) **(Figure 1)**. The Egger bias intercept test identified horizontal pleiotropy for the smoking-CAD pathway (intercept = 0.01, p = 0.046) **(Supplemental Table 3)**, although the findings remained robust in sensitivity analyses using MR methods that account for the possibility of horizontal pleiotropy **(Supplemental Figures 2-5)**, and when using an alternative genetic instrument for smoking (smoking initiation) **(Supplemental Figure 6)**.

**Figure 2:**
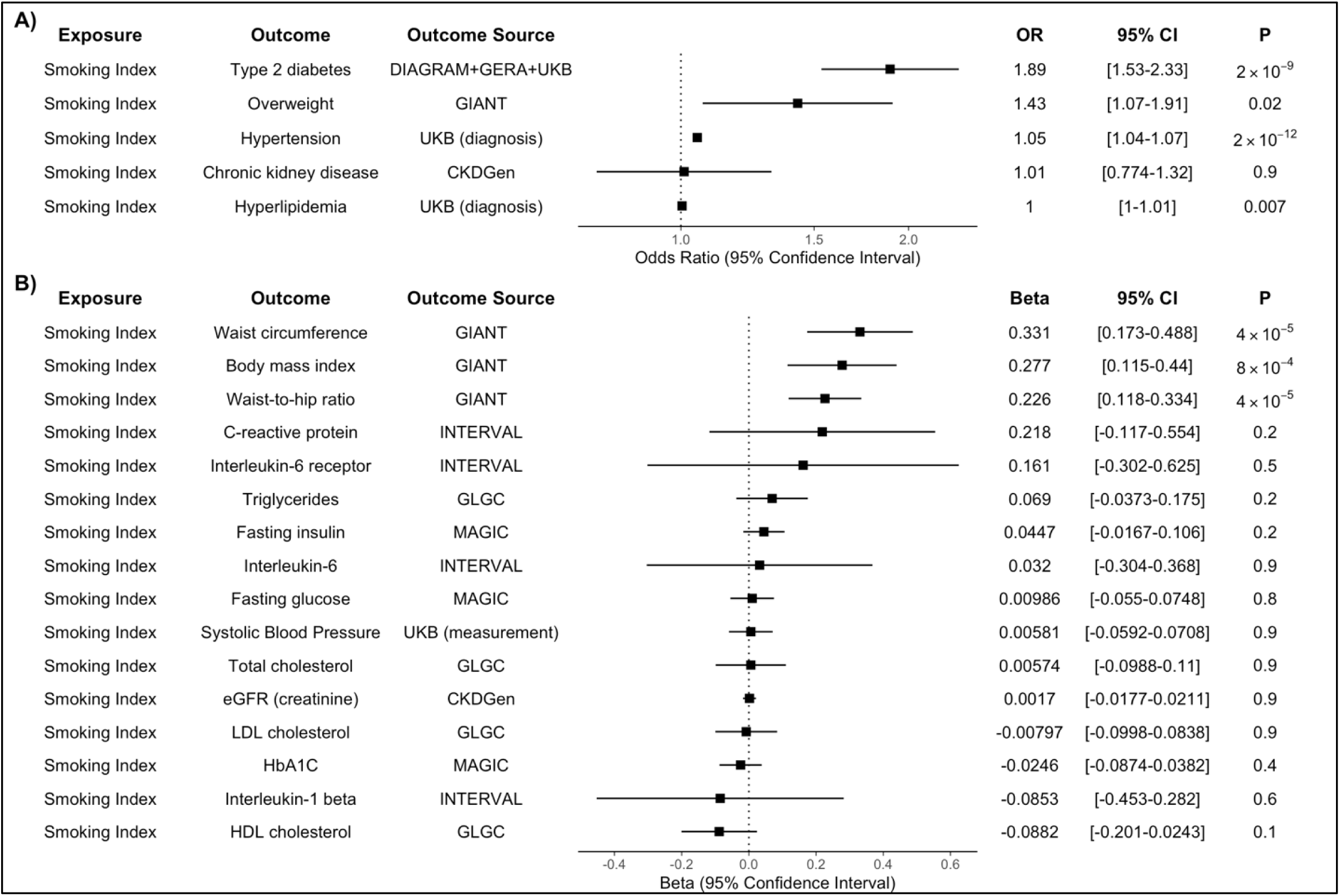
Total effect of smoking on risk of cardiometabolic risk factors for ASCVD. Inverse-variance weighted MR was performed to determine whether genetic liability to smoking altered risk of cardiometabolic risk factors for ASCVD. Genetic liability to smoking increased risk of both **(A)** binary traits, and **(B)** continuous traits that are common risk factors for cardiometabolic disease. Effect estimates are expressed per one standard deviation increase in lifetime smoking index. OR = Odds Ratio, CI = Confidence Interval.

We next determined if the causal effect of smoking differed between the ASCVD endpoints. The primary inverse-variance weighted effect estimate of smoking initiation on PAD was significantly greater than the estimates for large-artery stroke (OR_PAD_ 2.13 vs. OR_stroke_ 1.4; p_difference_ = 0.025) or CAD (OR_PAD_ 2.13 vs. OR_CAD_ 1.48; p_difference_ = 0.0041).

### Effect of Smoking on Cardiometabolic Risk Factors

We next considered the effect of increasing genetic liability to lifetime smoking on other cardiometabolic traits that are known ASCVD risk factors. Increasing genetic liability to lifetime smoking increased the risk of type 2 diabetes, hypertension, waist circumference, body mass index, and waist-to-hip ratio, with a suggestive (p < 0.05) increase in the risk of being overweight **(Figure 2)**. In sensitivity analysis considering a genetic instrument for smoking initiation as the exposure, results were similar **(Supplemental Figure 7)**. The MR Steiger testing confirmed the direction of these findings **(Supplemental Table 4)**.

### Cardiometabolic Risk Factors as Mediators of Smoking-ASCVD Risk

To evaluate whether increasing genetic liability to smoking increases risk of ASCVD directly, or whether the effect is mediated by traditional cardiovascular risk factors (type 2 diabetes, lipids, body mass index, and systolic blood pressure), we performed multivariable Mendelian randomization to estimate the direct effects of smoking. Increasing genetic liability to smoking increased risk of PAD, CAD, and stroke, after accounting for effects of smoking on each risk factor independently, and in a combined model considering all risk factors **(Figure 3)**. There was no significant attenuation of the effect estimates after accounting for traditional risk factors, or in a model further including interleukin-6 as a marker of inflammatory risk of ASCVD.

**Figure 3:**
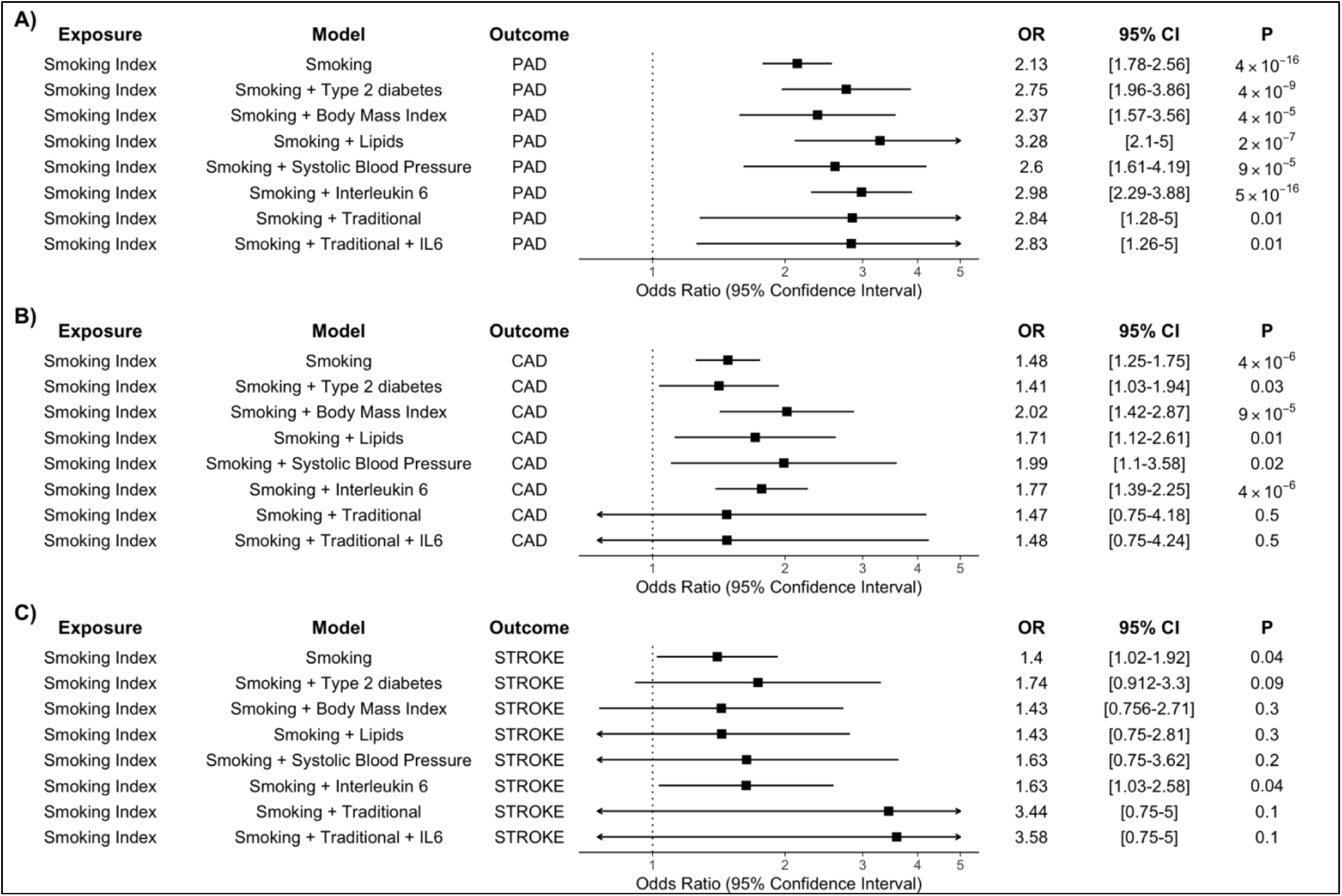
Direct effect of smoking on risk of PAD, CAD, and Stroke. Multivariable MR was performed to estimate the direct effect of smoking on ASCVD after accounting for the effects of smoking on other cardiovascular risk factors. The effect of smoking on PAD, CAD, and stroke was not substantially attenuated in models adjusting for traditional cardiovascular risk factors (Type 2 Diabetes, Body Mass Index, Lipids [LDL-C, HDL-C, Triglycerides], and Systolic Blood Pressure), or inflammation (IL6 levels). Odds ratios are expressed per one standard deviation increase in lifetime smoking index. OR = Odds Ratio, CI = Confidence Interval.

## DISCUSSION

Using Mendelian randomization, we leveraged population-scale human genetics to estimate the causal risk of smoking on cardiometabolic risk factors and atherosclerotic cardiovascular disease outcomes across diverse vascular beds (CAD, PAD, and stroke). By drawing from multiple large genome-wide association studies, we were able to consider an order-of-magnitude more cases of smoking, cardiometabolic risk factors, and ASCVD outcomes compared to previous observational studies. While observational studies remain at risk of bias due to residual confounding, the genetic instrumental variables utilized here in the Mendelian randomization framework provide effect estimates that are unconfounded from environmental factors. Because genetic variants are randomly assorted during meiosis, mimicking randomization in a clinical trial, we were able to estimate putative causal relationships between smoking and cardiometabolic traits.

Our results suggest that smoking has a direct atherogenic effect that varies across vascular beds. This finding is largely consistent with prior investigations of smoking on ASCVD. While the association between smoking and ASCVD has been demonstrated previously in observational studies, our Mendelian randomization analysis provides strong evidence consistent with a causal relationship. Our finding that smoking appears to more strongly influence the risk of PAD compared to CAD or stroke is consistent with recent results from the ARIC cohort, in which the effect of smoking was greatest for PAD.^4^ While the mechanism behind the stronger relationship between smoking and PAD is not clear, structural and functional differences within the vascular beds and the complex interplay between smoking and other ASCVD risk factors may contribute.^33,34^

The genetic liability to smoking is also associated with cardiometabolic traits that are themselves risk factors ASCVD. The MR finding that increasing genetic liability to smoking is associated with type 2 diabetes is consistent with recent observational and MR studies.^6,35–37^ We also identified increasing genetic liability to smoking as a risk factor for hypertension and increased waist circumference, body mass index, and waist-to-hip ratio, although prior studies have suggesting conflicting effects of smoking on these traits.^38–43^ A prior single-sample MR analysis from the Nord-Trøndelag Health Study (HUNT Study) found a protective effect of smoking on BMI, waist circumference, and hip circumference, but found no associations with blood pressure, lipids, or glucose levels.^40^ This study may have been limited by the single-sample design, modest study size, and weak single-SNP (rs1051730) instrument for smoking, which all may have contributed to bias toward observational estimates.^44^ A more recent MR study has identified a bidirectional relationship between smoking traits and BMI.^45^ Conflict among observational studies may be related to residual confounding or reverse causality. MR assumes that genetic variants proxying an exposure produce similar effects to the exposure itself, although this assumption may not always be valid. For example, lifetime exposure to adverse genetics may have different health consequences when compared to more concentrated environmental exposures, highlighted by the much larger protective effects of genetically lower LDL-cholesterol and systolic blood pressure on risk of coronary heart disease in comparison to effect estimates from randomized trials of treatments for these risk factors.^46,47^

The effect of increasing genetic liability to smoking on ASCVD outcomes appears to be independent from the effects of smoking on traditional cardiovascular risk factors. The point estimate of the direct effect of smoking (when jointly considering smoking and cardiometabolic risk factors) was similar (or greater) than the total effect, suggesting the possibility of causal interaction between smoking and traditional risk factors, which could be investigated using factorial MR in a single-sample setting.^48^ Proposed mechanisms by which smoking may independently contribute to cardiovascular events include hypercoagulability, increased myocardial work, decreased oxygen delivery (due to elevated carboxyhemoglobin levels), coronary vasoconstriction, and increased catecholamine levels, among others.^48^

The finding that smoking confers strong independent risk for ASCVD even when considering other traditional cardiovascular risk factors has important public health implications. More precise estimation of the effect of smoking on ASCVD outcomes may help calibrate the expected benefit of smoking cessation initiatives, and efforts to reduce the burden of cardiovascular disease should continue to focus on smoking cessation.

The current study must be interpreted within the context of its limitations. The study focused primarily on individuals of European ancestry, which may limit generalization to other populations, highlighting the need for genomic studies in diverse ancestral groups. The Mendelian randomization framework relies on a key assumption that the risk conferred by an exposure is equivalent whether mediated by genetics or environment, and that genetic risk is conferred through the exposure of interest rather than via pleiotropic effects ^12^. Although findings were consistent in sensitivity analyses using MR methods robust to the presence of pleiotropy, there may be gene-environment interactions, like those previously demonstrated at the *ADAMTS7* locus for CAD and the *CHRNA3* locus for PAD, that modify and alter the relationship between smoking and ASCVD outcomes ^9,49^. Although differences in the underlying structure of the ASCVD studies could affect the estimate of differential risk between the ASCVD outcomes, the two-sample Mendelian randomization framework tends to bias causal estimates toward the null, lending further confidence in our overall finding that smoking is strongly associated with increased risk of all ASCVD outcomes. Finally, future study of additional smoking-related traits, like duration/quantity of smoking and smoking cessation, and other MR methods may provide additional insight into potential differential effects of these traits in different vascular beds, clarifying recent observational findings that these traits may affect ASCVD risk.^4,50^

## CONCLUSION

Overall, increasing genetic liability to smoking increases risk of atherosclerotic cardiovascular diseases, with the strongest effect on peripheral artery disease, independent of traditional cardiovascular risk factors.

## Data Availability

The data that support the findings of this study are available from the corresponding author upon reasonable request.

## ACKNOWLEDGEMENTS

This research is based on data from the MVP, Office of Research and Development, Veterans Health Administration and was supported by award no. MVP000. This work was supported by US Department of Veterans Affairs grants IK2-CX001780 (SMD), and I01-BX003362 (PST). This publication does not represent the views of the Department of Veterans Affairs or the United States government. This work was supported by American Heart Association Strategically Focused Research Network in Vascular Disease grant 18SFRN33960373 (MSF); National Heart, Lung, and Blood Institute grants HL125032 (MSF) and R01HL142711 (PN); National Institute of Diabetes and Digestive and Kidney Diseases DK101478 and HG010067 (BFV); and a Linda Pechenik Montague Investigator Award (BFV). Data on coronary artery disease have been contributed by the CARDIoGRAMplusC4D and UK Biobank CardioMetabolic Consortium CHD working group who used the UK Biobank Resource (application number 9922). Data have been downloaded from www.CARDIOGRAMPLUSC4D.ORG. GWAS summary statistics for ischemic stroke have been contributed by the MEGASTROKE investigators. The MEGASTROKE project received funding from sources specified at http://www.megastroke.org/acknowledgements.html.

## ETHICAL APPROVAL

This study utilized only publicly available, deidentified summary statistics from previously published works, making it exempt from Institutional Review Board review at the University of Pennsylvania.

